# Foundation Model-Based Recommendation of Optimal Neoadjuvant Therapy in Breast Cancer

**DOI:** 10.1101/2025.10.03.25337255

**Authors:** Tuyen Vu, Ha X. Tran, Lin Liu, Jiuyong Li, Jia Tina Du, Thuc D. Le

## Abstract

Neoadjuvant therapy, involving treatment administered before surgery to shrink tumors, significantly impacts breast cancer management. However, current clinical approaches rely predominantly on limited clinical features, leading to suboptimal patient outcomes. To enhance therapeutic decision-making, we propose a novel foundation model-based recommendation framework (FDR) utilizing TabPFN, a deep learning model trained on extensive synthetic tabular data. Our method integrates multi-omics profiles with traditional clinical factors, enabling accurate counterfactual predictions for various drug combinations. Experimental results show that FDR markedly improves personalized treatment recommendations, resulting in a three-fold increase in recovery response rates. This study introduces the first multi-omics-informed neoadjuvant recommendation system, advancing precision oncology and demonstrating effectiveness even with limited patient data.

## 1 Introduction

Neoadjuvant therapy, defined as the administration of therapeutic agents before surgery, plays a crucial role in breast cancer management. ^1^ Its primary objectives include shrinking tumors to facilitate breast-conserving surgery and improving recovery outcomes. Current neoadjuvant regimens often combine chemotherapeutic agents such as taxanes and anthracyclines with targeted therapies like anti-HER2 agents. Historically, decisions regarding these regimens have relied on a limited set of clinical factors—including tumour size, grade, hormone receptor status and HER2 status—to guide drug selection and therapy planning.^2^ Clinicians commonly employ empirical risk stratification based on these criteria to develop neoadjuvant therapy plans. ^3^ However, this approach often leads to relatively low response rates, with only 26% of patients achieving a recovery response. ^4^ As shown in Figure 1, integrating machine learning into the breast cancer care process has the potential to support clinicians in treatment plan decisionmaking and improve recovery outcomes by offering data-driven insights.

**Figure 1:**
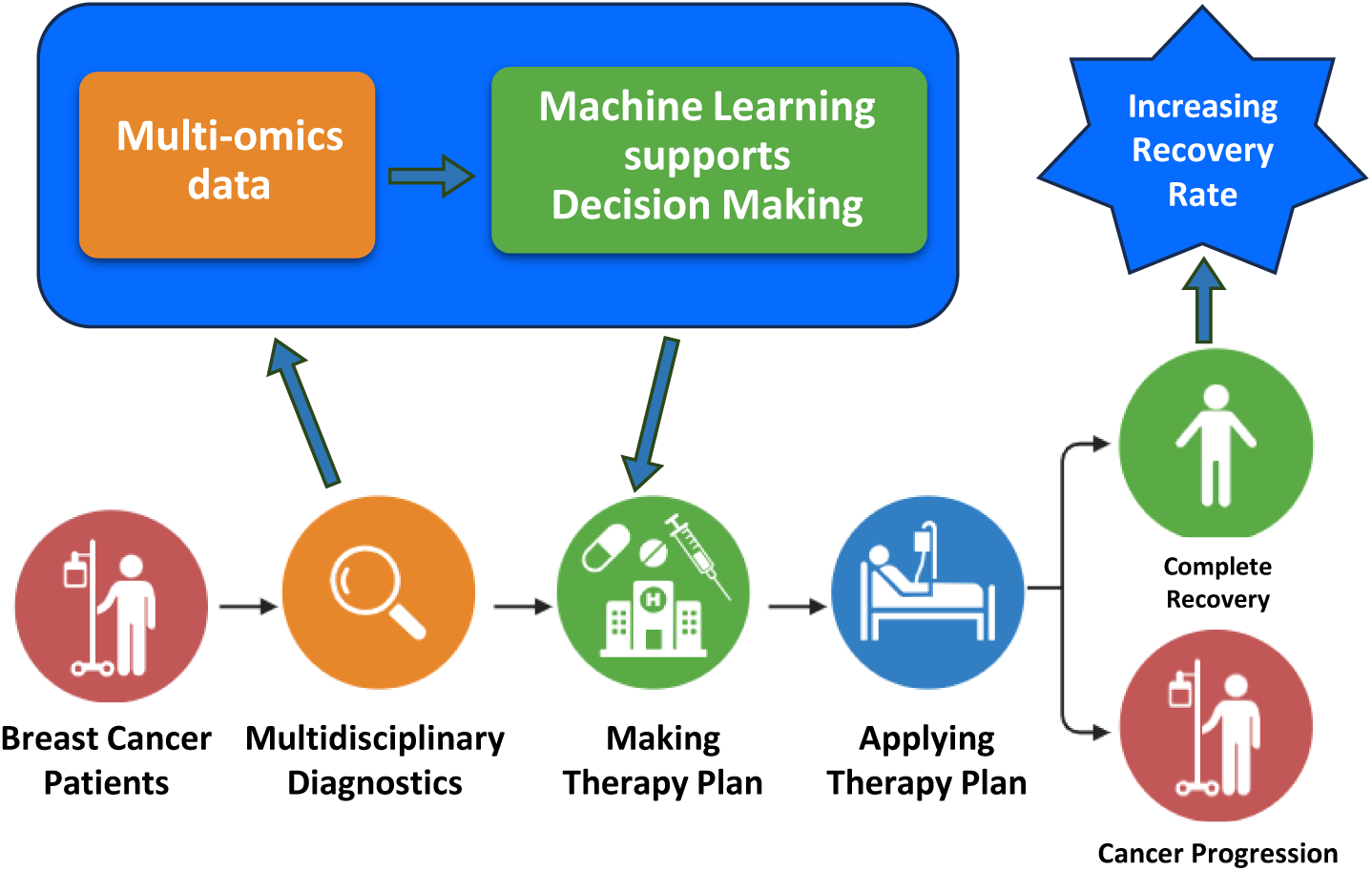
Breast Cancer Care Process with Machine Learning support for Treatment Decision-Making

This integration raises a fundamental question in personalized medicine: What is the optimal treatment (i.e., which drug combination) for each individual patient? Computational approaches aim to address the critical counterfactual question: What would the outcome have been if a different drug or drug combination had been selected for this patient? Addressing this question is challenging because only one outcome can be observed for each patient at any given time.

Possible solutions for answering that counterfactual questions are causal recommendation approaches.^5,6^ These methods focus on estimating heterogeneous treatment effects (HTEs)—the varying effects of treatments across distinct patient subpopulations. Subsequently, the features of new patients are mapped to these identified subpopulations, thereby recommending the optimal treatment based on estimated treatment effects.

Although causal recommendation approaches have demonstrated effectiveness in specific contexts, ^7–9^ they face several limitations. Firstly, they predominantly handle binary outcomes (e.g., recovered or not recovered) rather than utilising continuous treatment response metrics such as the Residual Cancer Burden (RCB) score. Secondly, these tree-based methods can be sensitive to training dataset size, as certain strata might lack sufficient cases to accurately estimate treatment effects. Lastly, their applicability is typically limited to datasets with a modest number of features, restricting their effectiveness in high-dimensional multi-omics datasets essential for accurate inference of patient treatment responses.

To bridge this gap, we utilise a foundation model for tabular data^10^ to recommend the optimal treatment for each individual based on their multi-omics profiles. Our approach first estimates counterfactual outcomes for various treatments (drug combinations) using the tabular foundation model and then recommends the optimal treatment for each patient based on these counterfactual predictions. The tabular foundation model, specifically TabPFN,^10^ is a deep learning architecture pre-trained on millions of synthetic tabular datasets. It employs prior knowledge captured from these diverse synthetic datasets, enabling it to generalize effectively and achieve high predictive accuracy across various real-world datasets. Its ability to rapidly estimate complex patterns makes it particularly suited for handling high-dimensional multi-omics data. In this paper, we leverage this foundation model’s capability to accurately predict counterfactual outcomes, enabling us to construct a robust recommendation system for personalized therapy selection. This work significantly contributes to precision oncology and improves breast cancer treatment outcomes through the following two key aspects:

1. **A New Foundation Model-based Recommendation Method for Personalized Medicine (FDR):** We propose a novel foundation model-based approach for therapy recommendation. By explicitly focusing on therapy regimens as the primary variable, our framework has demonstrated a three-fold increase in recovery response rates among patients who followed our recommendations compared to those who did not.
2. **Multi-omics-driven Recommendations:** We present the first study utilizing multi-omics data derived from tumor ecosystems to establish links between specific neoadjuvant therapy regimens and patient outcomes. This approach complements existing clinical practices by integrating molecular-level insights with traditional clinical features, offering a more comprehensive and precise framework for personalized therapy selection.

Experimental results demonstrate that the foundation model–based recommendation method, FDR, is effective for datasets containing diverse data types, including categorical and continuous features. Additionally, this method performs well even with small sample sizes, a common scenario in biomedical research, particularly for rare diseases. This capability addresses the limitations of traditional causality-based approaches, which typically require large datasets for training and are highly sensitive to hidden confounders. Moreover, FDR shows markedly better performance when applied to multi-omics data compared to using clinical features alone, achieving up to a 4-fold improvement in treatment recovery prediction. In out-of-distribution validation using independent real-world breast cancer datasets, FDR also demonstrates strong robustness, outperforming traditional baselines that fail under distributional shifts. These findings highlight FDR’s generalisability and effectiveness in complex, heterogeneous biomedical data environments.

## 2 Related Work

### 2.1 Neoadjuvant therapy response prediction

Machine learning approaches have shown promise in classifying patients likely to respond to certain treatments. ^4,11,12^ However, they typically rely on clinical features for prediction and do not incorporate specific therapy regimens as core model inputs. Instead, treatment is often treated as a fixed attribute or overlooked entirely, with models focusing solely on predicting outcomes based on patient characteristics.

Recently, with the increasing availability of multi-omics data and the growing sophistication of machine learning methods, numerous studies have focused on predicting treatment responses^4^ using multi-omics data. Approaches such as Multi-Omics Factor Analysis (MOFA) ^13^ integrate multi-omics data to uncover hidden interactions within the tumor ecosystem, while machine learning algorithms—including Support Vector Machines,^14,15^ Random Forests^16,17^ and LASSO regression^18^—identify biomarkers associated with therapy response. Additional network-based methods, such as Gene Set Enrichment Analysis (GSEA)^19^ and Cytoscape,^20^ elucidate disrupted biological pathways, aiding in the identification of factors driving therapy resistance or sensitivity.

Although these models are useful for predicting patient responses to neoadjuvant treatment in general, they fail to address the pivotal counterfactual question: What would the outcome have been if we had chosen a different drug or drug combination for this patient? Answering this question requires estimating counterfactual outcomes, as only one outcome can be observed for each patient at a time.

### 2.2 Non-Causal and Causal Recommendation Systems

Non-causality-based recommendation systems^21–23^ focus on generating recommendations to achieve optimal outcomes. The optimization is based on correlations between items, users, and outcomes, i.e., estimating. These models attempt to estimate under various modelling assumptions. If we consider intervention as the action of recommending items to users, represents pre-intervention associative relationships.

Causal recommendation systems (CRSs),^8,24,25^ by contrast, are interested in post-intervention effects caused by changes in a causal factor. An intervention that triggers the greatest causal effect on the outcome—and thus the greatest increase in the outcome—is recommended to users or patients. This approach aims to answer counterfactual what if questions for hypothetical scenarios: What would happen to a patient if we intervened on a factor or applied a treatment? By answering these questions, we can estimate the causal effects of different factors under a given intervention. This enables the recommendation of the most effective intervention or treatment to maximize the outcome for the patient.

In the context of this paper, CRSs aim to recommend the optimal neoadjuvant therapy plan for individual breast cancer patients. In contrast to traditional predictive approaches, these methods focus on estimating the causal effects of specific therapy combinations, thereby addressing the counterfactual question of which therapy would be most effective for each patient. Some CRSs build on the causal tree method by Athey and Imbens^5,7,26^ to estimate heterogeneous treatment effects (HTEs). By incorporating specific treatments as causal variables, these models can offer personalised recommendations that account for individual patient features. The novelty of this approach lies in integrating therapy plans directly into the causal model, thereby overcoming the limitations of general risk stratification.^3^ By quantifying how each treatment combination influences patient outcomes, we establish a data-driven basis for personalized therapy recommendations. Moreover, analyzing causal relationships in multi-omics data enables subgroup identification, facilitating more accurate estimations of treatment effectiveness for new patients.

## 3 Problem Definition

### 3.1 Neoadjuvant Therapy Plans for Breast Cancer

Neoadjuvant therapy typically consists of taxanes, anthracyclines and anti-HER2 agents. The chemotherapy regimen alternates between taxanes and anthracyclines over a median duration of 18 weeks (6 cycles). ^27,28^ In this study, neoadjuvant therapy refers to a combination of chemotherapy agents: taxane is administered in every cycle. Thus, two binary variables denote the presence or absence of anthracyclines and anti-HER2. Consequently, we can define four possible combination cases based on the number of drugs included in each regimen, encompassing all samples observed in the dataset. We denote these four drug combinations as four distinct Therapy Plans. For the observational data, if a patient receives a specific therapy plan *TP_k_*, we set *TP_k_* = 1, and the indicators for all other therapy plans are set to 0, as each patient receives only one therapy plan at a time. Table 1 summaries the Neoadjuvant Therapy Plan indicators.

**Table 1:** Neoadjuvant Therapy Plan Indicators.

| $TP_1$ | $TP_2$ | $TP_3$ | $TP_4$ | Drug Regimens |
| --- | --- | --- | --- | --- |
| 1 | 0 | 0 | 0 | Taxane, Anthracycline, anti-HER2 |
| 0 | 1 | 0 | 0 | Taxane and Anthracycline. |
| 0 | 0 | 1 | 0 | Taxane and anti-HER2 |
| 0 | 0 | 0 | 1 | only Taxane |

A Therapy Plan (TP) is defined as a specific combination of these agents. Taxanes are a key component of neoadjuvant therapy due to their efficacy in improving pathological complete response rates. ^29,30^

### 3.2 Clinical vs Multi-omics features

Table 2 presents examples of clinical and multi-omics features included in the multi-omics dataset used for breast cancer research. ^4^

**Table 2:**
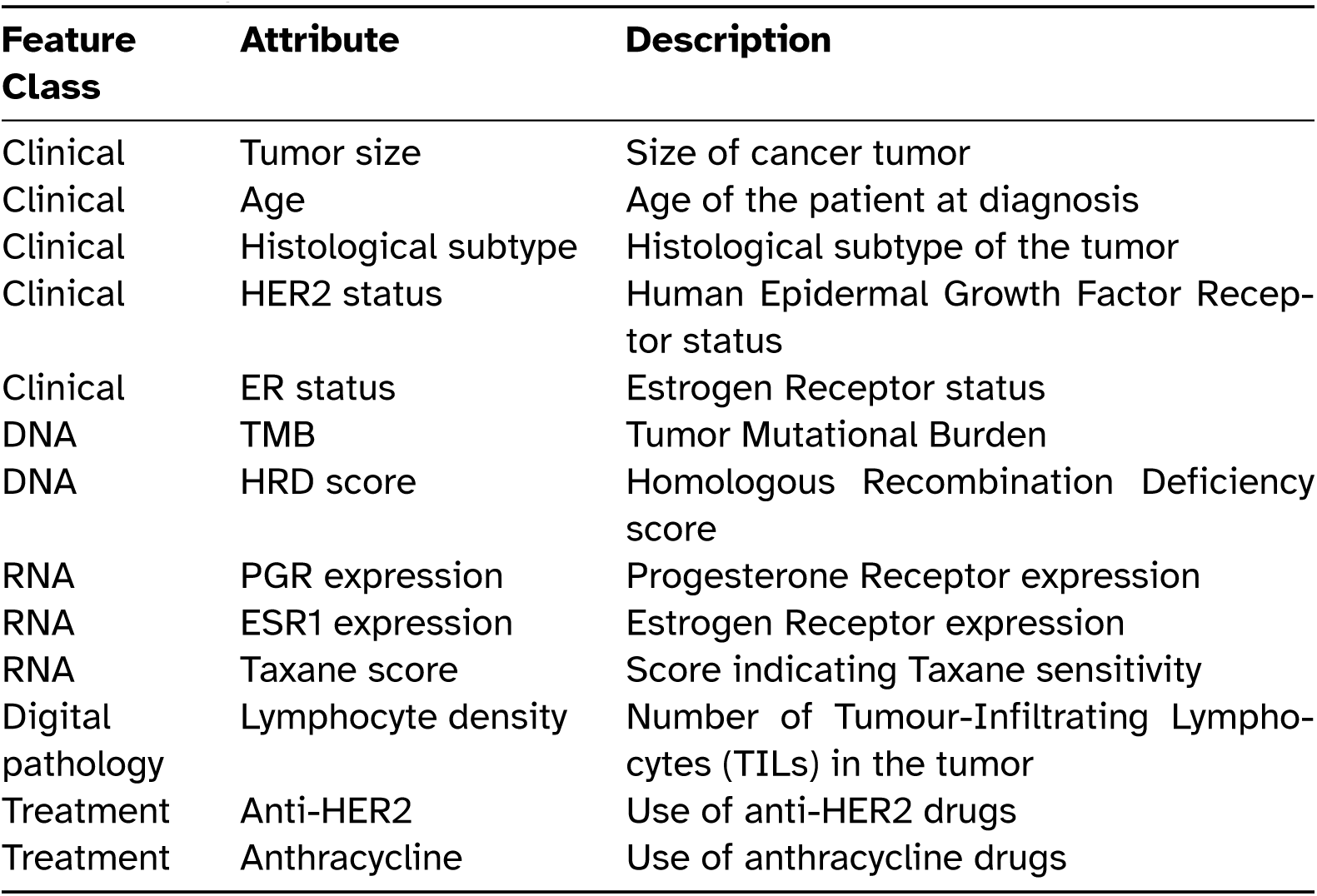
Example Features in the Multi-omics Dataset for Breast Cancer Patients.

| Feature Class | Attribute | Description |
| --- | --- | --- |
| Clinical | Tumor size | Size of cancer tumor |
| Clinical | Age | Age of the patient at diagnosis |
| Clinical | Histological subtype | Histological subtype of the tumor |
| Clinical | HER2 status | Human Epidermal Growth Factor Receptor status |
| Clinical | ER status | Estrogen Receptor status |
| DNA | TMB | Tumor Mutational Burden |
| DNA | HRD score | Homologous Recombination Deficiency score |
| RNA | PGR expression | Progesterone Receptor expression |
| RNA | ESR1 expression | Estrogen Receptor expression |
| RNA | Taxane score | Score indicating Taxane sensitivity |
| Digital pathology | Lymphocyte density | Number of Tumour-Infiltrating Lymphocytes (TILs) in the tumor |
| Treatment | Anti-HER2 | Use of anti-HER2 drugs |
| Treatment | Anthracycline | Use of anthracycline drugs |

Key clinical features include patient age, tumor size, histology, hormone receptor (ER and HER2) status, lymph node (LN) status, and ER Allred score. ^31^ Digital pathology, represented by the number of Tumour-Infiltrating Lymphocytes (TILs) in the tumor (Lymphocyte density), offers insights into immune micro-environments. ^4^ Given the complex biology of breast cancer, involving both malignant cells and their tumor microenvironment, ^32^ approaches that rely solely on broad clinical criteria are susceptible to suboptimal outcomes. ^4^ This underscores the critical need for more precise, patient-specific methods of planning neoadjuvant therapy—methods that extend beyond generalized clinical risk models and better account for the multifaceted nature of the tumor ecosystem.

Multi-omics data offer a promising route to address this need by providing a comprehensive view of tumor biology. ^11^ Tumors are heterogeneous ecosystems comprising malignant cells, stromal cells and immune components, all interacting within a dynamic microenvironment. ^33^ By integrating genomics, transcriptomics and other omics data, researchers can gain holistic insights into tumor cell characteristics, immune interactions and therapeutic responses.^4^ Advances in high-throughput techniques have further enabled the collection of vast multi-omics datasets, driving the development of novel analytical tools. ^34^ Leveraging these data provides clinicians with a deeper understanding of tumor biology and supports the design of more effective and personalized neoadjuvant therapy plans for breast cancer.

The treatment outcome is assessed using the Residual Cancer Burden (RCB) score, a widely used continuous metric that quantifies the amount of residual disease following neoadjuvant therapy in breast cancer patients. Developed by, ^27^ the RCB score integrates pathological measurements from the primary tumor and lymph nodes after surgery to estimate treatment response.^27^ The Residual Cancer Burden (RCB) score is a continuous metric used to quantify the amount of cancer remaining after neoadjuvant chemotherapy, offering a more nuanced assessment than binary outcomes. The calculation incorporates tumor size, cellularity, and lymph node involvement to reflect overall residual disease burden. Based on predefined cutpoints, the continuous RCB score is classified into four categories: RCB-0 (score = 0), indicating a pathological complete response (pCR); RCB-I (0 *<* score ≤ 1.36), representing minimal residual disease; RCB-II (1.36 *<* score ≤ 3.28), indicating moderate residual disease; and RCB-III (score *>* 3.28), representing extensive residual disease, corresponding to extensive residual disease. This stratification provides valuable clinical insight, allowing practitioners to interpret treatment effectiveness across a spectrum of response levels. ^35,36^ Compared to binary pCR endpoints, the RCB score offers a more nuanced and clinically informative assessment of treatment efficacy, making it particularly valuable for guiding post-surgical decisions.

### 3.3 Recommending the optimal neoadjuvant therapy for each patient

In this study, we use observational datasets with four drug combinations, denoted as mutually exclusive therapy plans (*TP*_1_, *TP*_2_, *TP*_3_, and *TP*_4_) as defined in Section 3.1. Each patient receives exactly one therapy plan, and the outcome is measured as a continuous variable (RCB score, where lower values indicate better outcomes). The objective is to recommend the optimal therapy plan for each patient by predicting treatment outcomes (RCB score) and selecting the plan with the lowest predicted value.

## 4 Methodology

### 4.1 Method overview

The proposed method consists of two main stages.

#### Stage 1: Obtaining Counterfactual Treatment Response Predictions

In this stage, a machine learning model (e.g., regression) or a foundation model (e.g., TabPFN) is trained on patient data to learn the relationship between input features (clinical, genomic, etc.) and treatment outcomes. The model is first fitted to a training dataset and then used to generate predictions on a counterfactual test dataset. These predictions represent the expected outcomes for each test patient under different therapy plans.

The ultimate goal is not just to predict outcomes under observed treatments, but to simulate counterfactual outcomes—that is, what would happen if each patient were assigned to a different therapy plan. To do this, the model generates separate outcome predictions for each possible therapy plan by altering the treatment indicator variables in the input data while keeping all other features constant. This simulates how each individual might respond to every available treatment option.

#### Stage 2: Personalized Recommendation

Once the model has generated counterfactual outcomes for all treatment options for each patient, the system identifies the optimal therapy plan— the one associated with the lowest predicted RCB score (i.e., the best outcome).

##### Algorithm 1 Foundation Model-Based Therapy Plan Recommendation (FDR)

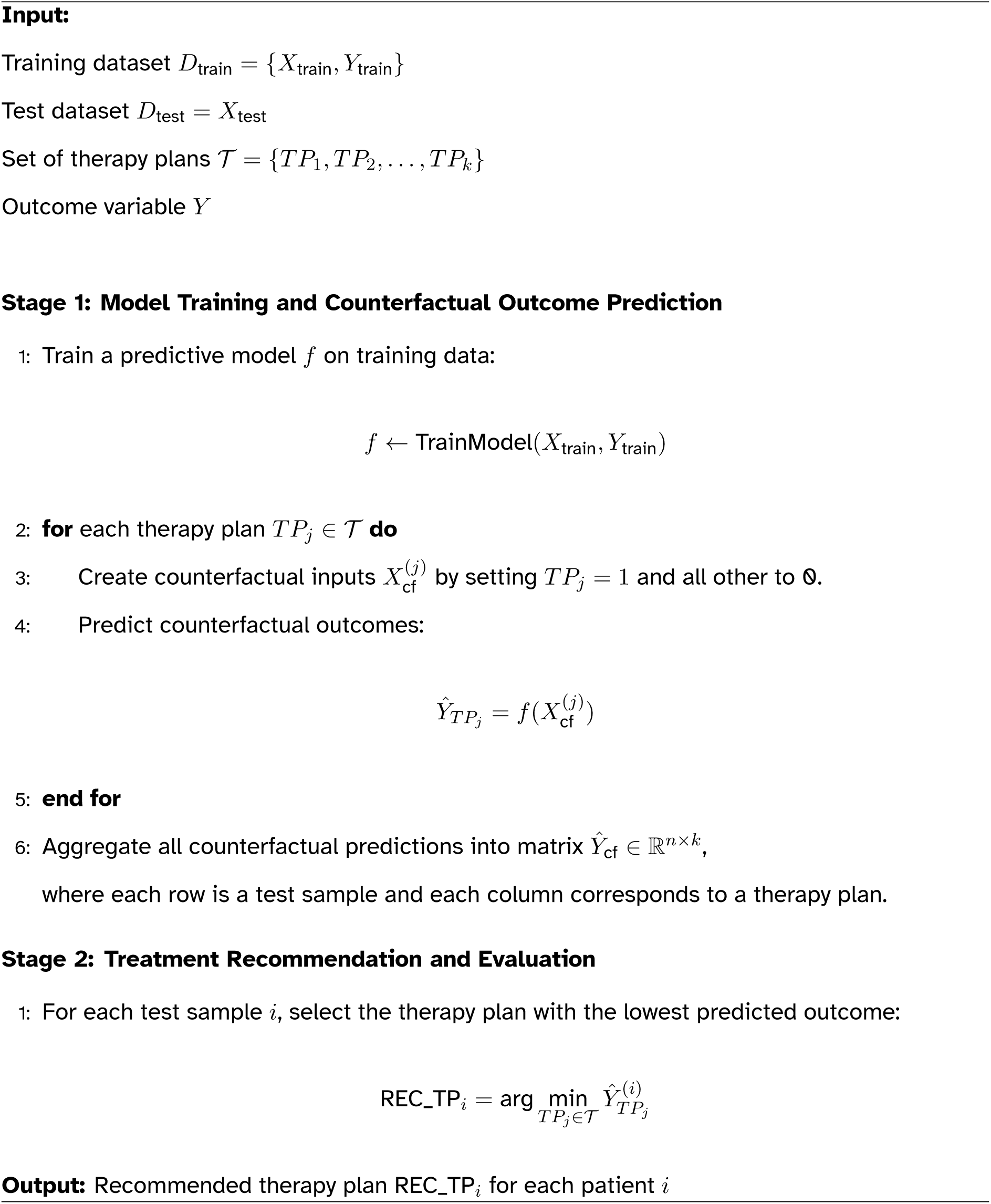

The FDR algorithm 1 outlines personalizes neoadjuvant therapy selection for breast cancer patients using foundation model-based method. It consists of two main phases: (1) Predictive Modelling and Outcome Estimation, and (2) Personalised Therapy Recommendation, which assigns the most effective treatment based on individual patient characteristics.

### 4.2 Predicting counterfactual outcomes of different Neoadjuvant Therapy plans

#### 4.2.1 Foundation model-based method

In this framework, we leverage TabPFN, a foundation model designed specifically for small-scale tabular data tasks, to estimate counterfactual outcomes for personalized treatment recommendation. Unlike conventional machine learning models that require explicit training on each new dataset, TabPFN employs a novel in-context learning mechanism. This approach processes the entire supervised learning task—comprising training examples, features, and test samples—as a single input sequence to a transformer model. Importantly, the model performs zero-shot prediction without any gradient-based updates, eliminating the need for re-training or hyperparameter tuning on new tasks.

The core of TabPFN is a Prior-Data Fitted Network (PFN), which approximates Bayesian inference over a distribution of tabular tasks. During its pretraining phase, the model was exposed to millions of synthetic classification tasks sampled from a rich, structured prior. These tasks simulate plausible real-world tabular problems, capturing causal relationships, low-dimensional manifolds, and other realistic data-generating patterns. As a result, TabPFN learns to implicitly perform posterior predictive inference by treating the observed data as in-context samples— effectively simulating how a Bayesian model would update its beliefs given new evidence.^10,37,38^

This inductive bias makes TabPFN exceptionally suited to counterfactual prediction. For each test sample, we can generate multiple counterfactual scenarios by modifying treatment indicators while keeping all other covariates fixed. TabPFN uses the provided context of training data and modified test instances to estimate the expected outcome under each hypothetical treatment. This approach is especially powerful in low-data regimes, as the model’s reasoning is guided by strong structural priors learned during pretraining, rather than requiring large training datasets to generalize.

#### 4.2.2 Traditional machine learning methods

Traditional machine learning methods—such as regression—are adapted to estimate counterfactual treatment outcomes by following a two-stage approach similar to the FDR framework. In the first stage, a predictive model is trained on observed patient data—including clinical, genomic, and treatment information—to learn the relationship between features and treatment outcomes. After training, the model is applied to new patient data to estimate potential outcomes under different therapy scenarios. To simulate counterfactuals, the model generates predictions under each possible therapy plan by modifying the treatment variables while keeping other inputs unchanged. This process estimates how each patient would respond to all available treatments. The personalized recommendation is then made by selecting the therapy plan associated with the most favorable predicted outcome.

#### 4.2.3 Causality-based method

CausalTree is a causality-based approach that differs from other baseline models, as it is specifically designed to estimate the effect of a therapy plan on the outcome. It is suitable for binary outcomes and requires a sufficiently large sample size to construct reliable trees.

Our goal is to provide personalized therapy plan recommendations tailored to individual patients. We recognize that the effectiveness of a given therapy plan can vary significantly from one patient to another, leading to heterogeneity in treatment responses. This necessitates the estimation of the Conditional Average Treatment Effect (*CATE*) for a binary treatment-therapy plan *TP*, which represents the *ITE* and helps identify the most effective therapy plan. To achieve unbiased CATE estimation for a specific subgroup, we can apply inverse probability of treatment weighting (IPTW) using propensity score,^39^ as follows:

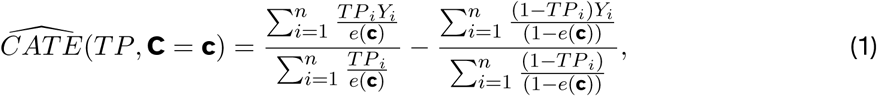

where *e*(**c**) = *P* (*TP* = 1|**C** = **c**) is the propensity score, ^40^ the probability of therapy plan assignment. The score can be estimated using logistic regression. In this study, the covariate set **C** corresponds to the set **X**.

The objective of the method extends beyond estimating the average treatment effect in observational data. Its primary aim is to identify patient subgroups that exhibit diverse treatment effects. Rather than focusing solely on estimating the average treatment effect across the entire population, the CausalTree method seeks to uncover subgroups with varying conditional treatment effects and to estimate the heterogeneous treatment effects of neoadjuvant therapy plans with interpretability. ^5^

### 4.3 Personalized therapy recommendations

Once the model has generated simulated outcomes for all treatment options for each patient, the system identifies the optimal therapy plan, the one associated with the lowest predicted adverse outcome. The recommended treatment is then compared with the actual treatment received by each patient, using the concepts of *Following* and *Not Following* to facilitate the evaluation of the model’s recommendation performance.

## 5 Experiments

### 5.1 Experiment settings

#### 5.1.1 Datasets

In this study, we utilised observational data from the TransNEO neoadjuvant breast cancer clinical trial^4^ and the ARTemis and Personalized Breast Cancer Programme (PBCP) studies. ^41^ The TransNEO trial initially recruited 180 women, of whom 147 cases with complete molecular and digital pathology information were included in our analysis. The second dataset comprised an external cohort of 75 patients from the ARTemis studies, who also received neoadjuvant therapy. ^41^

The data comprised clinical, digital pathology, genomic and transcriptomic information derived from pre-treatment biopsies of breast tumors in patients who underwent neoadjuvant therapy. Each patient was assigned an individualised therapy plan involving a combination of three drugs: taxane, anthracycline and anti-HER2, with the outcome measured by the continuous Residual Cancer Burden (RCB) score and categorical RCB class.^27^ A rising RCB score indicates a growing burden of remaining disease following neoadjuvant therapy and a rise in chemoresistance.^4^

We also used 2 public clinical datasets for breast cancer research from the GEO (Gene Expression Omnibus) data repository hosted by NCBI (National Center for Biotechnology Information) including GSE25066^42,43^ and GSE41998.^44^ GSE25066 comprises clinical profiles from 508 breast cancer patients who underwent neoadjuvant taxane-anthracycline chemotherapy. This dataset^42^ includes clinical annotations such as estrogen receptor (ER), progesterone receptor (PR), and HER2 status, as well as treatment outcomes like pathologic complete response (pCR) and residual disease (RD). GSE41998 contains gene expression data from a randomized, open-label, involving early-stage breast cancer patients treated with sequential neoadjuvant therapy: four cycles of doxorubicin and cyclophosphamide (AC), followed by either ixabepilone or paclitaxel. ^44^ The dataset includes pre-treatment tumor biopsies and is instrumental in analyzing biomarkers associated with response to these chemotherapeutic agents. Both datasets are instrumental in advancing our understanding of breast cancer biology and treatment response.^45^

- **TransNEO Clinical Dataset (clin_TransNEO):** Clinical data from the TransNEO trial, ^4^ comprising 147 samples with 8 clinical features, treatment arms, and RCB outcomes.
- **ARTemis Clinical Dataset (clin_ARTemis):** Clinical data from the ARTemis and PBCP studies,^41^ including 72 samples with 8 clinical features, treatment arms, and RCB outcomes.
- **GSE25066 Clinical Dataset (clin_GSE25066):**^42^ We used 116 samples from this dataset that have complete treatment arm information and treatment outcomes. Each sample includes 14 clinical features, 2 treatment arm indicators, a binary outcome (pCR), and a categorical outcome (RCB.category).
- **GSE41998 Clinical Dataset (clin_GSE41998):**^44^ We used 265 samples from this dataset. Each sample includes 9 clinical features, 2 treatment arm indicators, a binary outcome (pCR), and a categorical outcome (RCB.category).
- **TransNEO Multi-omics Dataset (multi_TransNEO):** Data from the TransNEO neoadjuvant breast cancer trial. ^4^ This dataset includes 147 samples with 74 multi-omics features and binary, categorical, and continuous RCB outcomes.
- **ARTemis Multi-omics Dataset (multi_ARTemis):** Data from the ARTemis and Personalized Breast Cancer Programme (PBCP) studies.^41^ The dataset originally contains 75 samples; however, we used 72 samples that have complete RCB outcome data, including 74 multi-omics features.
- **Combined TransNEO and ARTemis Multi-omics Dataset (multi_Trans_ART):** A merged dataset combining TransNEO and ARTemis data, resulting in 219 samples with 74 multi-omics features, treatment arms, and RCB outcomes.

#### 5.1.2 Baselines

From the literature review, no existing method specifically focuses on therapy plan recommendation; current machine learning approaches primarily rely on traditional methods for predicting treatment responses. We present the first study utilising multi-omics data for therapy plan recommendation. To enable comparison, we developed baseline recommendation models to evaluate the performance of the proposed foundation model-based recommendation method, FDR.

##### Traditional Machine Learning models for recommendation

To implement the baseline models for treatment recommendation tasks, seven widely used machine learning methods are employed: CatBoost, Neural Network, Linear Regression, Random Forest, Support Vector Regression (SVR), and XGBoost, among others. Some of these models (Random Forest, Support Vector Machine) were previously applied in multi-omics predictive studies,^4^

##### Causality-based model for recommendation

In this study, the CausalTree method could not be applied to the ARTemis, GSE25066, and GSE41998 datasets due to their limited sample sizes and imbalanced treatment distributions. To implement the CausalTree model for treatment recommendation, separate trees are constructed for each therapy plan, treating the plan as the intervention and pathological complete response (pCR) as the binary outcome. The CausalTree algorithm partitions patients into subgroups and estimates the Conditional Average Treatment Effect (CATE) within each leaf, representing groups with similar treatment responses. Four Causal Trees are built—each corresponding to a different therapy plan—to estimate the treatment effect of that plan. A personalised recommendation is then made for each patient by selecting the therapy plan with the highest estimated CATE, as this plan is expected to offer the greatest potential for recovery.

- **CT (CausalTree):**^5,26^ builds a binary regression tree model in two stages, but focuses on estimating heterogeneous causal effect.
- **CB (CatBoost):**^46^ an algorithm for gradient boosting on decision trees.
- **NN (Neural Network):** ^47^ Using the MLPRegressor module from the scikit-learn library. ^48^ MLPRegressor is Multi-Layer Perceptron, an old term for a neural network.
- **LR (Linear Regression):**^49^ Using the LinearRegression module from the scikit-learn library. ^48^
- **RF (Random Forest):**^16^ Using the RandomForestRegressor module from the scikit-learn library. ^48^
- **SVR (Support Vector Regression):**^14^ Using the SVR module from the scikit-learn library. ^48^
- **XGB (XGBoost):**^50^ XGBoost uses gradient boosted decision trees, a supervised learning algorithm that combines weak learners to create a strong learner.

To align with the proposed FDR approach for recommending the optimal therapy plan for breast cancer patients, the baseline methods were used to predict the counterfactual outcome RCB score. The therapy plan associated with the lowest predicted outcome RCB score was then recommended.

#### 5.1.3 Evaluation Metrics

To assess the effectiveness of our proposed FDR model in therapy recommendation, we define two evaluation metrics: **Coverage-Adjusted Uplift** and **Average RCB Score** between the *Following* and *Not Following* groups. A recommendation is classified as *Following* if the actual treatment received by the patient matches the model’s recommended therapy plan. Conversely, a case is labeled *Not Following* if the patient’s treatment differs from the model’s recommendation. These two groups allow us to evaluate whether patients who follow the model’s guidance tend to achieve better clinical outcomes. The **Coverage-Adjusted Uplift** measures the relative recovery rate by the number of patients who have a positive outcome between the groups, while the **Average RCB Score**, using metrics such as the Residual Cancer Burden (RCB) score, quantifies the difference in treatment response. This comparison helps determine whether adherence to the recommended therapy plan is associated with improved patient recovery.

##### Normalising for Coverage: Coverage-Adjusted Uplift (CAU)

When evaluating treatment recommendation policies, it is not sufficient to compare the recovery rate of patients who follow the recommendation against those who do not. A policy may achieve a high uplift within the matched (“Following”) group, but if it only applies to a very small proportion of the overall population, its impact on clinical outcomes is limited. Conversely, a policy with moderate uplift but broader applicability can generate greater benefit overall. To make policies directly comparable, the uplift must therefore be *normalised for coverage*.

Let *p*_rec|follow_ denote the recovery probability among patients who followed the recommended treatment, and *p*_rec|not_ denote the recovery probability among those who did not. Let #follow be the number of patients who followed the recommendation and *N* the total number of patients. The **coverage-adjusted uplift (CAU)** is defined as:

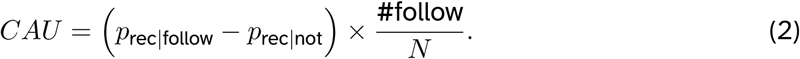

The first term, *p*_rec|follow_ − *p*_rec|not_ captures the difference in recovery rates between patients who follow the recommendation and those who do not, i.e. the per-patient uplift. The second term, #follow/*N*, represents the coverage, the fraction of patients for whom the policy provides a recommendation. Multiplying these terms yields the expected improvement in recovery probability at the population level.

In practice, the CAU can be interpreted as the number of additional recoveries (per patient, or scaled to per 100 patients) attributable to the recommendation policy, taking into account both its effectiveness and its coverage. This makes CAU a fair and interpretable metric for comparing different treatment recommendation strategies.

##### Comparison of Average RCB Score between Following and Not Following Groups

We utilised bar plots to visualise the number of recovered patients across different machine learning models, comparing two patient subgroups: those who followed the model’s recommended therapy plan and those who did not. Each method is represented by a pair of bars, bars with a “+” hatch for the Following group and bars with a “/” hatch for the Not Following group, with sample sizes annotated above each bar.

#### 5.1.4 Implementation

Our method is implemented using Python with the TabPFN model from TabPFN package ^1^. The baselines are implemented using Python with Random Forest, Support Vector Machine and Neural Network models from scikit-learn package ^2^, CatBoost package ^3^, and XGBoost package ^4^. Codes and public datasets are available for public access at GitHub ^5^.

### 5.2 Results

The results of the experiment show the comparison of all methods in terms of metrics: Coverage-Adjusted Uplift and the Average RCB Score.

#### Coverage-Adjusted Uplift Comparison

The results in Table 3 report the CAU achieved by different treatment recommendation methods across multiple clinical and multi-omics datasets. CAU normalises the uplift in recovery rates by the proportion of patients covered by the recommendation policy, thus reflecting the overall improvement in recoveries at the population level. Higher CAU values indicate more effective policies that not only achieve a larger difference in recovery between Following and Not Following groups, but also apply to a broader fraction of patients.

**Table 3:** CAU Comparison for all Methods in all Datasets.

| Dataset | FDR | CT | CB | NN | LR | RF | SVR | XGB |
| --- | --- | --- | --- | --- | --- | --- | --- | --- |
| TransNEO clinical | **3.98%** | -1.32% | -4.03% | 2.04% | -2.72% | -0.82% | 2.93% | 1.11% |
| ARTEMIS clinical | -1.92% | NA | 6.83% | 6.83% | 2.50% | **7.93%** | 6.83% | 7.40% |
| GSE41998 clinical | 6.51% | NA | **20.26%** | -5.61% | 1.55% | 5.92% | -5.07% | -0.73% |
| GSE25066 clinical | 0.86% | NA | **7.55%** | 0.07% | -2.12% | 4.44% | -2.79% | 2.18% |
| TransNEO multi-omics | **12.00%** | 4.11% | 10.12% | -0.71% | 2.52% | -0.66% | 2.93% | 3.63% |
| ARTEMIS multi-omics | **7.40%** | NA | 4.15% | -0.06% | -13.31% | -2.99% | 6.83% | 6.83% |
| Combined TransNEO + ARTEMIS multi-omics CV | **16.68%** | 5.30% | 8.84% | 0.10% | -5.55% | 4.84% | 4.84% | 3.49% |
| TransNEO and ARTEMIS multi-omics OOD | **11.39%** | 1.90% | 10.39% | 0.24% | -21.76% | 0.97% | 8.21% | 0.53% |

**Abbreviations:** CAU: Coverage-Adjusted Uplift, CT: Causal Tree, CB: CatBoost, NN: Neural Network, LR: Linear Regression, RF: Random Forest, SVR: Support Vector Regression, XGB: XGBoost, CV: Cross Validation, OOD: Out Of Distribution, NA: Not Available (Causal Tree could not be applied due to limited sample sizes or imbalanced treatment distributions).

The proposed FDR method consistently outperforms and is the best-performing method across all multi-omics datasets. For instance, in the combined TransNEO and ARTemis multi-omics dataset under the cross-validation setting, FDR achieved a CAU of 16.68%, the highest among all compared methods. Similarly, in the out-of-distribution evaluation, training on TransNEO and testing on ARTemis, FDR achieved a CAU of 11.39%, substantially higher than competing models. These results demonstrate that patients following the recommendations of FDR were substantially more likely to experience favourable outcomes compared to those who did not.

A key observation from Table 3 is that incorporating multi-omics data markedly improves the performance of treatment recommendation models compared to relying on clinical data alone. Across both the TransNEO and ARTemis datasets, most methods exhibit limited or inconsistent gains when restricted to clinical features, but achieve substantial improvements when multi-omics features are included. Notably, FDR improves its CAU from 3.98% (clinical only) to 12.00% (multi-omics) on TransNEO, and from −1.92% (clinical only) to 7.40% (multi-omics) on ARTemis. This trend highlights the importance of integrating genomic and transcriptomic features alongside clinical variables to provide more accurate and robust treatment recommendations.

Overall, the superior CAU performance of FDR across both cross-validation and OOD settings illustrates its strong potential for guiding personalised chemotherapy decisions. By effectively leveraging multi-omics data and accounting for patient heterogeneity, FDR maximises the expected recovery benefit at the population level while remaining clinically interpretable.

#### Out-of-Distribution (OOD) Validation

We further evaluated model robustness in an OOD setting, where models were trained on the TransNEO dataset and tested on an independent dataset, ARTemis. The results demonstrate that FDR maintains strong performance under distribution shift, achieving a CAU of 11.39%. In contrast, most baseline methods exhibited substantial degradation, with CAU values close to zero or even negative, indicating little to no improvement when following their recommendations. For example, CT and RF achieved CAU values of only 1.90% and 0.97%, respectively, while methods such as LR and SVR produced negative uplifts. Only CatBoost reached a competitive positive value (10.39%), but it still lagged behind FDR.

These results highlight the advantage of using pretrained foundation models in scenarios with unknown or shifted data distributions. By leveraging multi-omics features and accounting for heterogeneity, FDR generalises more effectively than traditional machine learning baselines, thereby providing more reliable treatment recommendations in realistic clinical settings where patient populations differ across cohorts.

#### Comparison of Average Continuous RCB scores

Among all the models evaluated, the FDR model shows the most significant difference in RCB scores between the Following and Not Following subgroups as the Figure 2. The Patients who followed the FDR recommendations had a notably lower median RCB score compared to those who did not, suggesting a strong association between adhering to FDR, guided therapy and improved treatment outcomes. This large gap indicates the model’s effectiveness in recommending optimal therapy plans that may contribute to better pathological response and a higher likelihood of recovery. In contrast, baseline models show smaller or inconsistent differences between the subgroups.

**Figure 2:**
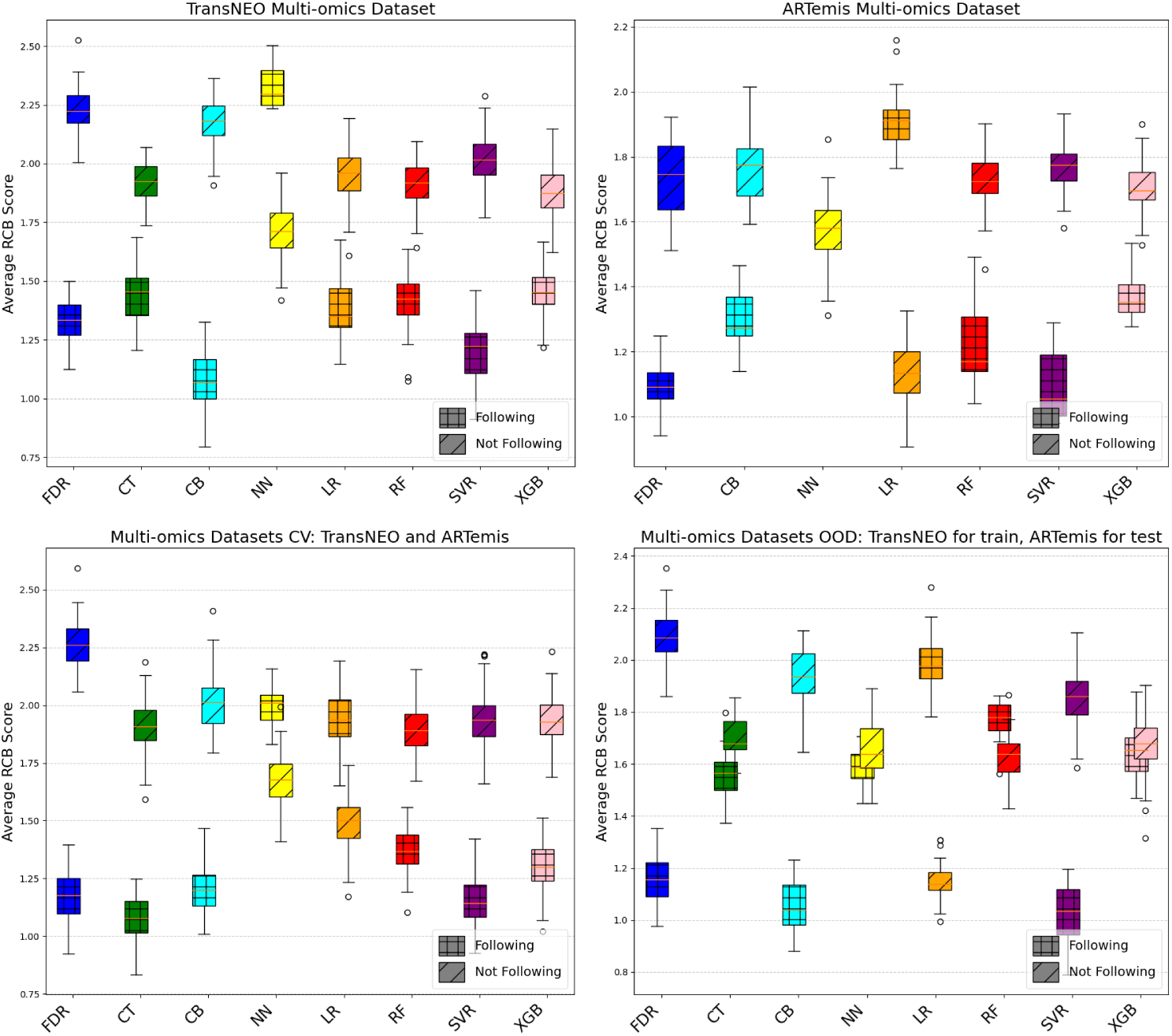
Comparison of Average RCB Score between Following and Not Following groups across Methods using Multi-omics datasets

Overall, this analysis highlights that the FDR model not only produces accurate predictions but also provides actionable recommendations that, when followed, are linked to significantly improved clinical outcomes.

## 6 Conclusions

In conclusion, this research introduces a novel approach for recommending personalised neoadjuvant therapy plans for breast cancer patients, addressing the limitations of traditional clinical risk stratification methods. The proposed FDR model, which incorporates multi-omics data from clinical, digital pathology, genomic and transcriptomic sources, offers a more effective and data-driven method for determining optimal treatment options. By applying foundation model-based techniques, the FDR model predicts the counterfactual outcomes of different therapy plans and recommends the one most likely to lead to a fully recovery.

This personalized recommendation system has the potential to transform breast cancer treatment by providing more tailored and effective plans for individual patients. The findings highlight the importance of integrating multi-omics data and foundation model-based methods into clinical decision-making, ultimately enhancing the precision and effectiveness of neoadjuvant therapies for breast cancer. The proposed model is not intended to replace healthcare providers or biologists, but rather to serve as a promising tool to support more informed decision-making. It represents a shift from traditional clinical guidelines toward a more data-driven, patient-specific approach. Future research could expand this framework to incorporate survival data, enabling the assessment of long-term treatment impacts and further refining therapy personalization to improve patients’ overall outcomes.

## Data Availability

All data produced in the present study are available upon reasonable request to the authors

https://ega-archive.org/studies/EGAS00001004582

https://www.ncbi.nlm.nih.gov/geo/query/acc.cgi?acc=gse41998

https://www.ncbi.nlm.nih.gov/geo/query/acc.cgi?acc=gse25066

## Footnotes

1 https://github.com/PriorLabs/TabPFN

2 https://scikit-learn.org/

3 https://pypi.org/project/catboost/

4 https://pypi.org/project/xgboost/

5 https://github.com/vntuyen/fdr

